# Influence of ACEi and ARB use on HAI Response to Seasonal Influenza Vaccination

**DOI:** 10.1101/2025.08.12.25333523

**Authors:** Frank N. Bunks, Ted M. Ross

## Abstract

**Background:** Angiotensin Converting Enzyme Inhibitors (ACEi) and Angiotensin Receptor Blockers (ARB) have anti-inflammatory properties via decreasing AT1R binding/levels, reducing Reactive Oxygen Species (ROS) and cytokine activation. This study investigated whether these medications can negatively impact hemagglutination inhibition (HAI) responses to influenza vaccination.

**Methods:** Participants, over the age of 18, were consented and enrolled in the study to receive an influenza vaccine during the 2024-2025 influenza season. Participants were classified into individuals on either ACEi or ARB. Healthy controls were selected based on age-sex-body mass index (BMI) matching and a second control group of participants were diagnosed with hypertension (HTN). but on taking these medications. Analysis focused on relative risk ratios (RR) for seroconversion to the influenza vaccine, in addition to day 28 geometric mean titers (GMT), number of seroprotected participants, and fold change was carried out.

**Results:** We observed a lower trend of seroconversion amongst participants taking either medication compared to the HTN controls and did not observe any differences compared to the healthy controls. There were no differences in day 28 GMT, although the HTN controls had statistically significant higher fold changes in HAI titers compared to the healthy controls. Compared to the treatment groups, the HTN controls had a non-significant higher fold change against all three strains included in the influenza vaccine.

**Conclusions:** Overall, the use of medications did not impact seroconversion when compared to healthy controls, but participants did have a lower trend for seroconversion compared to the HTN controls. This could be due to reduced inflammatory markers in people taking these medications, but this reduction in titer is similar to that in healthy participants. More studies comparing inflammatory markers related to medications in people are needed. In addition, determining the impact of the use of ACEi and ARB on lymphocyte responses is critical for effective influenza vaccination strategies.

## Introduction

Hypertension (HTN) is a cardiovascular disease characterized by increased blood pressure and inflammatory state [1]. In HTN, the renin enzyme converts the hormone angiotensinogen to angiotensin I, which is then converted to angiotensin II, via Angiotensin Converting Enzyme I (ACE1). Angiotensin II, in turn, binds to the angiotensin II type 1 receptor (AT1R), the main receptor responsible for increasing both blood pressure and inflammation [2]. This inflammatory cascade puts the person, especially those greater than 65 years old, at increased risk of mortality, hospitalization, ICU admission, and 30-day readmission when infected with viruses such as Influenza [3-5].

Angiotensin Converting Enzyme Inhibitors (ACEi) and Angiotensin Receptor Blockers (ARB), prescribed to over 40 million people in the United States, are common medication to treat HTN [6, 7]. ACEi inhibit the actions of Angiotensin Converting Enzyme 1 (ACE1), a vasoconstrictor/pro-inflammatory enzyme responsible for converting angiotensin I to angiotensin II [8]. On the other hand, ARBs block the binding of Angiotensin II to the Angiotensin II receptor Type 1 (AT1R) [9]. These medications lead to a subsequent decrease in binding to AT1R through decrease in angiotensin II levels, for ACEi, and displacement of angiotensin II binding, for ARB, resulting in reduced reactive oxygen species (ROS), cytokine activation, sympathetic nervous system (SNS) activation, AT1R levels, and NF-κB activation [10-12].

ACEi/ARB use is linked to reduced morbidity and mortality from influenza, including lower 30-day mortality, fewer ICU admissions, and a decreased risk of viral complications. [13-16]. These effects were attributed to an increase in ACE2 caused by the medications, which counteracts ACE1, decreasing inflammation through anti-inflammatory mechanisms such as increased Angiotensin (1-7) and lower inflammatory cytokine activation [17, 18]. These qualities may also negatively impact immune responses following influenza vaccination. The impact of ACEi/ARB on vaccination is limited, however, cardiovascular medications, such as statins that have immunosuppressive effects, reduced vaccine effectiveness in these populations compared to healthy people administered the same vaccines [19].

To address the gap in the current research, patients taking either ACEi or ARB were selected from a larger cohort of patients from the University of Georgia (UGA) trial [20] and compared against a healthy and non-healthy control group to determine if there is any significant effect on influenza vaccine responses. To consider vaccine effectiveness, seroconversion rates, antibody geometric mean titers (GMT), antibody fold changes, and post-vaccination seroprotection rates were examined. Overall, the use of ACEi or ARB did not have any effect on hemagglutination inhibition assay (HAI) response to influenza vaccination when compared with healthy controls, but these medications can decrease influenza vaccine seroconversion rates compared to people with HTN not on either ACEi or ARB.

## Methods

### Cohort Enrollment

All study procedures involving human participants were reviewed and approved by the WIRB-Copernicus Group Institutional Review Board (WCG IRB #20224877 for the influenza vaccine studies at the University of Georgia, as well as by the University of Georgia Institutional Review Board. Participants for the influenza vaccination study were enrolled during the 2024– 2025 season. All participants were enrolled in Athens, Georgia, USA. Informed consent forms were distributed, and written consent was obtained from all individuals prior to their inclusion in the studies. Study procedures included the collection of demographic data, and the collection of serum samples.

Eligible participants were adults aged 18 years and older and received tetravalent inactivated influenza (TIV) Fluzone vaccine (Sanofi Pasteur, Swiftwater, PA, USA), FLUCELVAX (Seqirus Inc, Holly Springs, NC, USA), FLUAD (Seqirus Inc, Holly Springs, NC, USA), FLUMIST (AstraZeneca, Cambridge, UK) and FluBlok (Sanofi Pasteur, Swiftwater, PA, USA). The egg based vaccines protected against A/Victoria/4897/2022 (H1N1), A/Thailand/8/2022) (H3N2), and B/Austria/1359417/2021 (B/Victoria). Meanwhile, the cell based vaccines protect against A/Wisconsin/67/2022 (H1N1), A/Massachusetts/18/2022 (H3N2), and B/Austria/1359417/2021 (B/Victoria)[21]. Data collection included demographics, age, sex, body mass index (BMI), and race/ethnicity with PBMCs collected on day 0 pre-vaccination and day 28 post-vaccination. Four sub-groups were established from this larger study participants; participants that reported the use of ACEi, participants that reported the use of ARB, participants reported HTN without the use of ARB or ACEi, and selected healthy controls. The healthy controls were selected through matching participants based on age, sex, and BMI. The outline of this process is shown in Figure 1.

**Figure 1.**
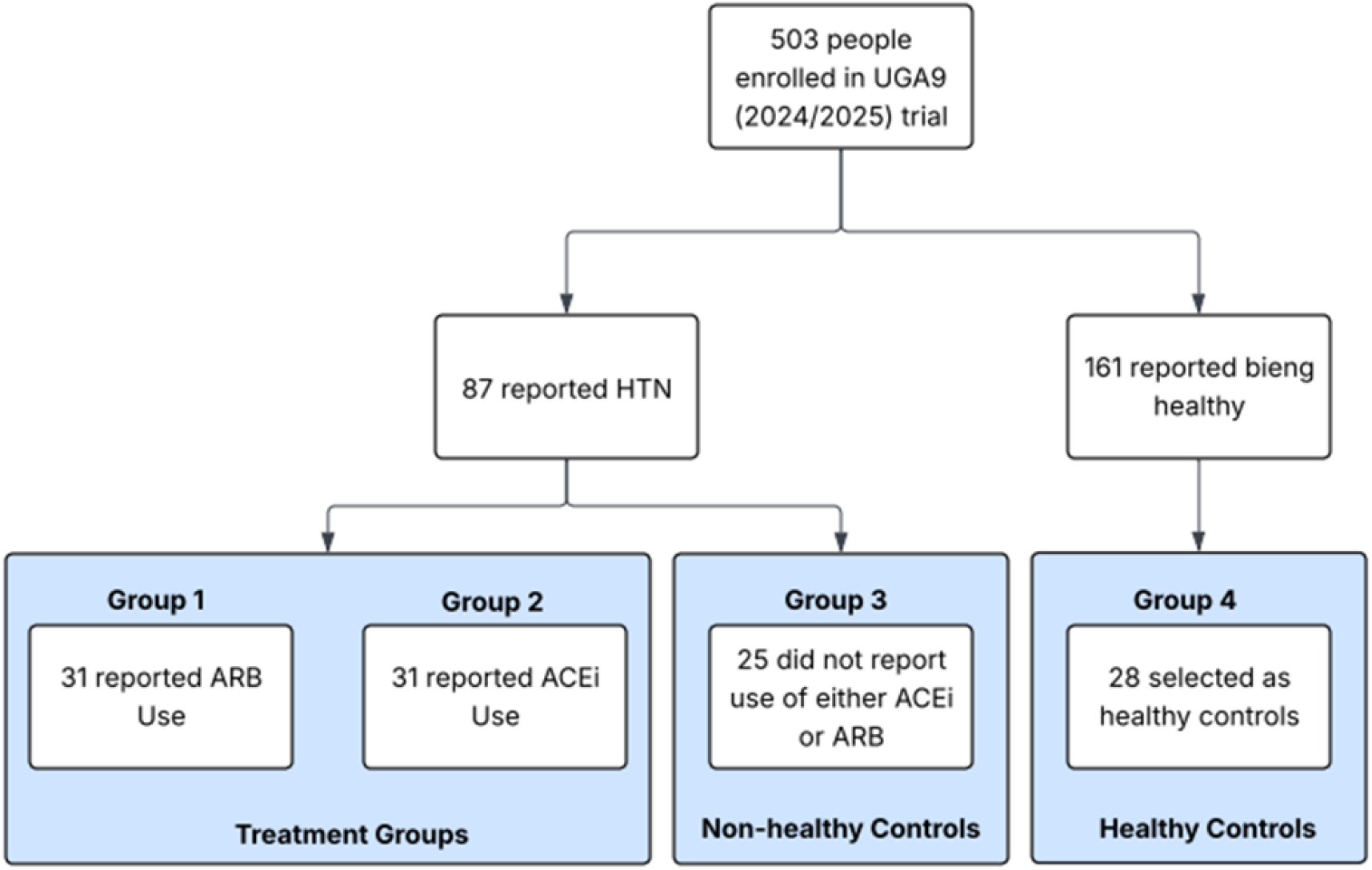
Outline of Groups compared in this study. Participants were enrolled during 2024/2025 Influenza season to the University of Georgia (UGA) 9 trial. They were then broken down into two initial groups based on whether they reported having Hypertension or whether they reported being healthy. They were then broken down into 4 groups based on the use of Angiotensin Receptor Blockers, Angiotensin Converting Enzyme, whether they had hypertension but did not report use of either medication, or if they were an age-sex-bmi matched healthy control.

### Hemagglutination Inhibition (HAI) Assay

Sera was aliquoted in tubes and treated with 3 volumes of receptor destroying enzyme (RDE) in 1x phosphate buffered saline (PBS) and then incubated overnight at 37 □C. Following incubation, the RDE with sera was heat inactivated in 56 □C for 30-45 minutes, and the sera was allowed to cool to RT and 6 volumes of PBS was added to make a sera dilution of 1:10. In a 96 V-bottom well microplate, 50 µL of PBS and 50µL influenza virus was added and serially diluted in a two-fold dilution. 50 µL of red blood cells (turkey blood for H1N1 and Victoria, and guinea pig blood for H3N2), and then gently mix and incubate at room temperature for 30 min. The plates were tilted to read the last agglutinated well and virus at 1:8 HA working concentration were prepared. For the assay, 25µL of PBS were added to wells 2 through 11 and with 50µL of PBS added to well 12. Then 50µL of RDE sera was added to the first well and serially diluted in a two-fold dilution, in addition 25µL of 1:8 working virus was added to all of the wells, except for column 12. Finally, the plate was mixed, incubated at room temperature for 20 min, 50µL of 0.8% RBC was added and further incubated at room temperature for 30 min. To read the results, plates were tilted at a 45 degree angle for ∼20 seconds.

### Endpoints and Statistical Analysis

The primary endpoint in this study was the Relative Risk (RR) of seroconversion, defined as ≥4-fold HAI increase from day 0 to day 28 with ≥1:40 HAI at day 28. Secondary endpoints in this study are geometric mean titers (GMT), fold changes from day 0 and day 28, and number of people who are seroprotective, defined as having a day 28 ≥ 40 HAI. Statistical significance was done using Fishers exact tests for the RRs and number of seroprotective people, and Kruskal-Wallis t-tests were done for the GMT and fold changes. In this study, significance was defined as having a p value ≤0.05 (^*^ = p≤0.05, ^**^ = p≤0.01, ^***^ = p≤0.001, ^****^ = p≤0.0001).

## Results

### Demographics of Participants

503 people were enrolled in the UGA trial during the 2024-2025 influenza season. Of these 503 participants, 87 reported having HTN and 161 reported having no comorbidities and on no medication. 31 people reported taking ARB and were put into the first treatment group, 31 reported taking ACEi and were put into the second treatment group. Furthermore, 25 people that had HTN did not report the use of ARB and/or ACE and were put into the non-healthy control group. Finally, 28 people from the 161 healthy participants were selected as controls for the treatment groups through age-sex-BMI matching (Figure 1).

The average age across the four groups was 67 (64-68) with a BMI of 29.67, 47% males and 53% females. The breakdown of each cohort demographic is demonstrated in table 1. Furthermore, comparisons of age and BMI showing the similarity between the treatment groups and their controls is shown in supplemental figure 1. While vaccine type is not an endpoint or consideration in our study, 56% of participants received either FLUZONE Standard or High Dose, 27% received FLUAD, 12% received FLUBLOK, 6% received FLUCELVAX, and 1% received FLUMIST.

**Table 1:** Demographics of participants included in this study.

| Cohort | Female (n, %) | Male (n, %) | Age (Average) | BMI (Average) |
| --- | --- | --- | --- | --- |
| ACE Inhibitor (n = 31) | 16 (51.6%) | 15 (48.4%) | 68 | 30.12 |
| Angiotensin Receptor Blocker (n = 31) | 16 (51.6%) | 15 (48.4%) | 68 | 31.07 |
| Non-Healthy Controls (n = 25) | 14 (56.0%) | 11 (44.0%) | 64 | 29.98 |
| Healthy Controls (n = 28) | 15 (53.6%) | 13 (46.4%) | 67 | 27.36 |

### Effect of Medication Usage on Seroconversion

Unadjusted comparisons, between the 87 participants reporting HTN and the 161 healthy participants were analyzed for influenza vaccine seroconversion by HAI assay (Figure 1) are shown in Figure 2. Participants with HTN have increased RR values against H1N1 (1.429, 95% CI [0.936 – 2.325]) and H3N2 (1.470, 95% CI [1.119 – 2.916]). However, against B/Victoria, people with HTN had decreased RR values compared to healthy participants (0.9302, 95% CI [0.6170 – 1.374]). The RR seen in the unadjusted analysis was only significant for H3N2 (Figure 2). There were no differences observed in the day 28 GMT between the HTN and healthy participants in H1N1 (20.33 vs 20.44; p=0.9745), H3N2 (53.04 vs 57.56; p=0.5735), B/Victoria (68.09 vs 53.94; p=0.0616). On the other hand, significant increase in fold change for people with HTN were observed for H1N1 (2.568 vs 2.088; p = 0.0436) and H3N2 (4.151 vs 3.957; p = 0.0134), but not for B/Victoria (2.275 vs 2.389; p = 0.9360).

**Figure 2.**
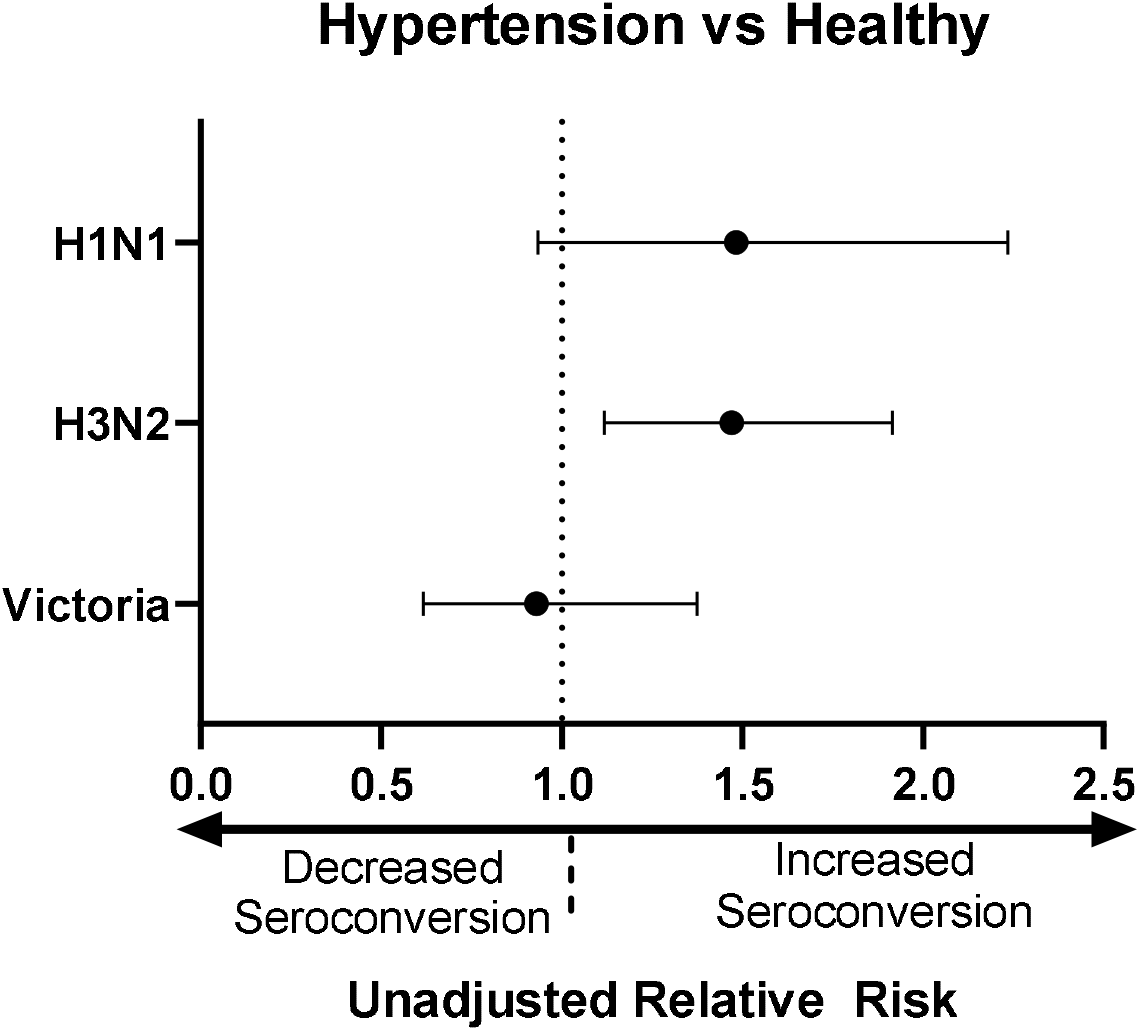
Forest plots depict the unadjusted Relative Risks Ratio with 95% Confidence Interval [CI] of HAI seroconversion in participants with hypertension (n=87) compared to people who are healthy (n=161). RR values greater than 1 favors HTN as the “risk” of seroconversion increases. RR values less than 1 favor the control as the “risk” of seroconversion decreases. RR values equal to 1 favor neither HTN nor control as there is no difference in seroconversion “risk”.

Upon adjusting for use of ACEi/ARB as well as participant age, sex, and BMI (Figure 1), no significance difference was observed on seroconversion outcomes from the use of ACEi/ARB (Figure 3). Although, it should be noted that there were consistent trends for lower RR when comparing the treatment groups to the non-healthy controls, as well as comparing the non-healthy controls to the healthy controls (Figure 3a-c) (Table 2). These trends were most notable in the H3N2 strain for treatment vs non-healthy (0.8065, 95% CI [0.4434 – 1.2672] and 0.6019, 95% CI [0.3635 – 0.9587]) and H1N1 for non-healthy vs healthy (1.68, 95% CI [0.7185 – 4.023]) (Figure 3a-b) (Table 2). There was no observable trend for higher or lower RR seroconversion compared to healthy controls across all three strains since there were increases against H3N2 in participants using ACEi, decreases against B/Victoria. There were also decreases against H3N2 and B/Victoria in participants with ARB (Figure 3a-c and Table 2). Although not statistically significant, the RR for seroconversion was higher in non-healthy controls compared to healthy controls across all strains; H1N1 (1.68, 95% CI [0.7185 - 4.023]), H3N2 (1.26, 95% CI [0.8357 – 1.93]), B/Victoria (1.008, 95% CI [0.4591 – 2.052]) (Figure 3a-b) (Table 2).

**Table 2:** Summarizes the RR ratio values with 95% Confidence Interval [CI] in the forest plot (Figure 1) and their p values demonstrated with Fishers Exact test.

|  | H1N1 |  | H3N2 |  | Victoria |  |
| --- | --- | --- | --- | --- | --- | --- |
|  | RR (95% CI) | P | RR (95% CI) | P | RR (95% CI) | P |
| ACEi vs Healthy | 1.204 (0.4931 - 2.99) | 0.7659 | 1.016 (0.6516 - 1.605) | >0.999 | 0.7226 (0.334 - 1.545) | 0.5721 |
| ARB vs Healthy | 1.204 (0.4931 - 2.99) | 0.7659 | 0.7583 (0.4434 - 1.272) | 0.4309 | 0.7467 (0.3457 - 1.592) | 0.5729 |
| ACEi vs Non-healthy | 0.7168 (0.3327 - 1.567) | 0.5599 | 0.8065 (0.5371 - 1.204) | 0.401 | 0.7168 (0.3277 - 1.567) | 0.5599 |
| ARB vs Non-healthy | 0.7168 (0.3327 - 1.567) | 0.5599 | 0.6019 (0.3635 - 0.9587) | 0.0552 | 0.7407 (0.3391 - 1.615) | 0.5616 |
| Non-healthy vs Healthy | 1.68 (0.7185 - 4.023) | 0.3604 | 1.26 (0.8357 - 1.93) | 0.39 | 1.008 (0.4891 - 2.052) | >0.999 |

**Figure 3.**
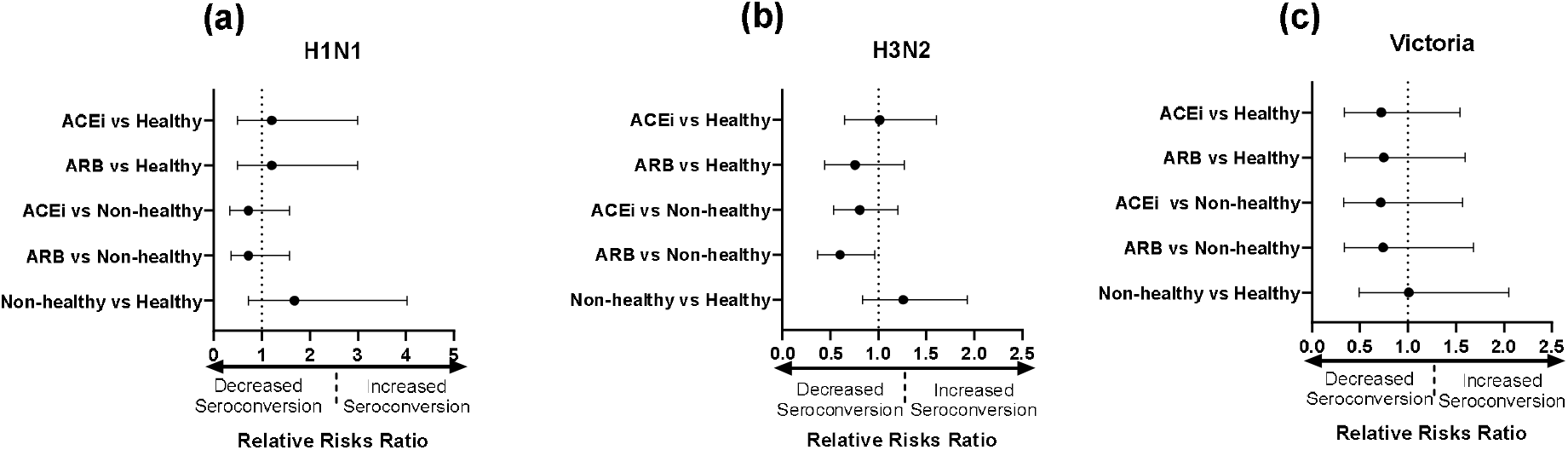
Forest plots showing the Relative Risks Ratio with 95% Confidence Interval [CI] of Seroconversion with comparisons between the treatment vs control groups, and non-healthy vs healthy control groups. The forest plots are separated based on virus strain: (a) forest plot for H1N1 (b) forest plot for H3N2 (c) forest plot for Victoria. RR values greater than 1 favor the use of the medication as the “risk” of seroconversion increases. RR values less than 1 favor the control as the “risk” of seroconversion decreases. RR values equal to 1 favor neither HTN nor control as there is no difference in seroconversion “risk”.

### Medication Influence on Overall HAI Response

Similarly, to the seroconversion response, no statistically significant difference was observed in GMT and seroprotection. All cohorts averaged HAI titers >1:40 HAI on day 28 against H3N2 and B/Victoria, but had lower HAI titers against the H1N1 vaccine component (Table 3), matching the pattern observed in the HAI seroconversion results (Table 3). Across all strains, the non-healthy controls had the highest geometric mean fold changes, with a 4-fold increase against H3N2. The non-healthy controls had a statistically significant increase in the H3N2 fold changes compared to the healthy controls (4.333 vs 2.394, p = 0.0298) (Figure 4e) (Table 3). There was no difference in the fold change in HAI titers observed between the in participants taking different medications, as well as both the non-healthy controls and healthy controls.

**Table 3:** Summary of the Endpoints used in this study per each cohort across the three strains: H1N1, H3N2, and B/Victoria.

|  | H1N1 |  |  |  | H3N2 |  |  |  | Victoria |  |  |  |
| --- | --- | --- | --- | --- | --- | --- | --- | --- | --- | --- | --- | --- |
|  | GMT | SP (n%) | SC (n%) | Fold Change | GMT | SP (n%) | SC (n%) | Fold Change | GMT | SP (n%) | SC (n%) | Fold Change |
| ACEi (n=31) | 17.49 | 11 (35.4%) | 8 (25.8%) | 2.297 | 48.92 | 21 (67.7%) | 18 (58%) | 3.891 | 69.96 | 23 (74.2%) | 8 (26%) | 2.639 |
| ARB (n=31) | 20.95 | 13 (42%) | 8 (25.8%) | 2.144 | 56.57 | 21 (67.7%) | 13 (41%) | 3.403 | 72.94 | 27 (87%) | 8 (26%) | 1.954 |
| Non-healthy (n=25) | 23.62 | 10 (40%) | 9 (36%) | 3.595 | 54.26 | 18 (72%) | 18 (72%) | 4.333 | 60.63 | 21 (84%) | 9 (36%) | 2.285 |
| Healthy (n=28) | 14.14 | 8 (29.5%) | 6 (21.4%) | 1.852 | 52.52 | 20 (71.4%) | 16 (57.14%) | 2.394 | 80 | 23 (82%) | 10 (35.7%) | 2.394 |

Abbreviations: GMT, Geometric Mean Titer; SP, Seroprotection; SC, Seroconversion

**Figure 4.**
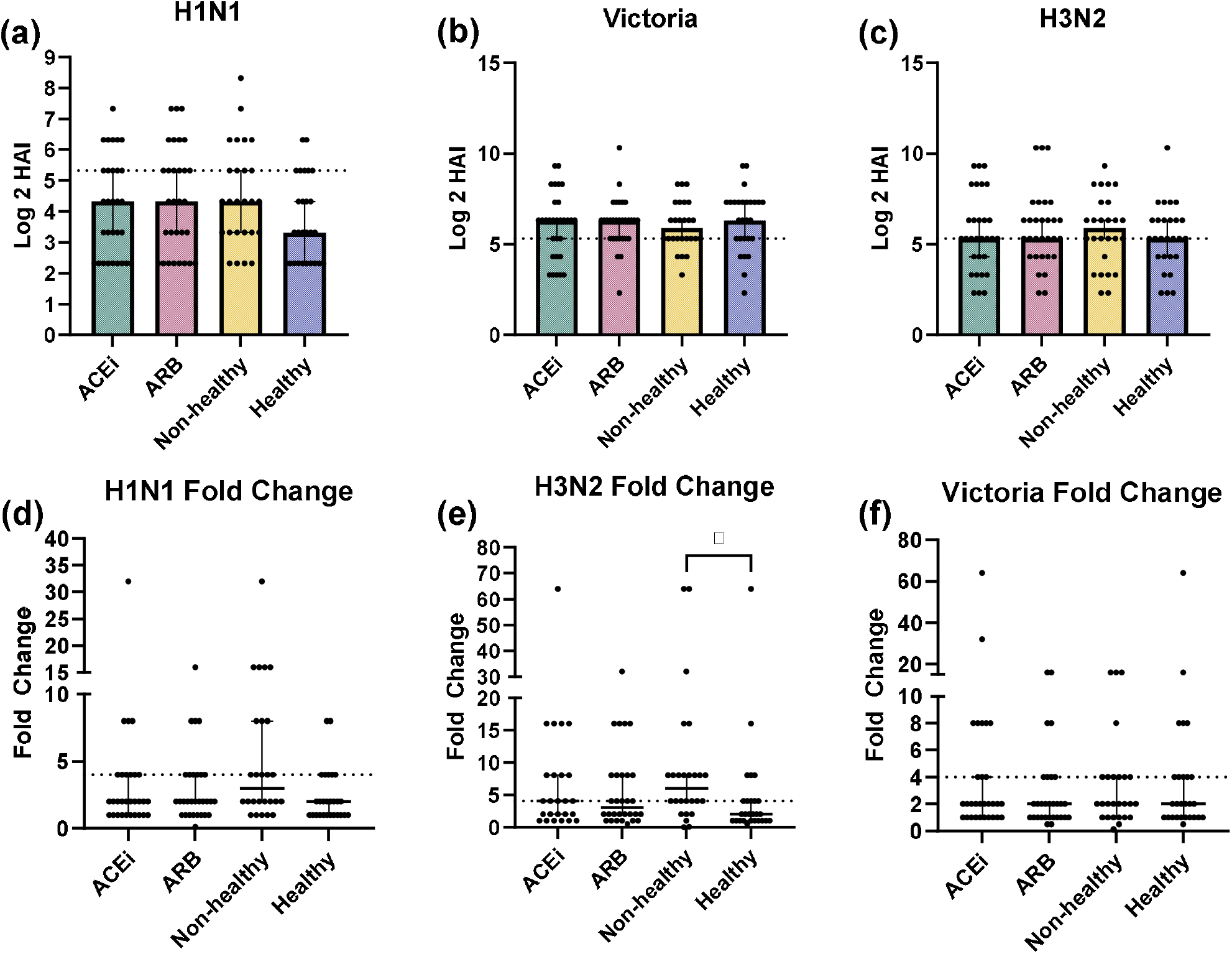
Day 28 HAI titers (median with 95% CI) by each participant group against the H1N1 (a), H3N2 (b), and B/Victoria (c). Fold change between the HAI titer at day 28 compared to day 0 against the H1N1 (d), H3N2 (e), and B/Victoria (f). Statistical significance was determined using Kruskal-Wallis test.

## Discussion

In this case-control study, participants were administered one of six 2024-2025 seasonal influenza vaccines. Serum samples were collected at day 0 (pre-vaccination) and day 28 (post-vaccination) were tested for HAI activity against influenza viruses matching the components of the vaccine. Participants with HTN and healthy people and observed increased RR values for H1N1 and H3N2 (significant for H3N2), and decreased RR values for B/Victoria. Adjustments for medication usage (ACEi and ARB), as well as matching individual age, sex, and BMI in healthy controls was performed. In this adjusted analysis, there were non-significant trends of lower RR from participants using ACEi and ARB to the non-healthy control and healthy control participants. In addition, there were higher, but not statistically different, RR seroconversion HAI titers from non-healthy control participants compared to healthy control participants. There were no differences in seroprotection or HAI GMT between groups, however, the non-healthy control participants had statistically significant higher fold changes in HAI titers compared to the healthy control participants against H3N2 component of the vaccine. HTN presents an exaggerated inflammatory state through increased inflammation from factors such as AT1R binding, IL-6, Il-1β, and ROS activation [22].

The use of ACEi can significantly reduce C-reactive protein (CRP), interleukin (IL-6), IL-8, and monocyte chemoattractant protein-1 [23, 24]. Similarly, ARB significantly decrease IL-6, neutrophil counts, IL-1β, CRP, and C-X-C motif chemokine ligand 1 (CXCL1) [23, 25, 26]. Furthermore, the use of these medications can also diminish the amount of damage associated molecular patterns and neoantigens released from endothelial cells during vasoconstriction. Comparisons between the inflammatory markers/cells present in people on medications and healthy controls is lacking. The decrease in inflammation resultant from the medications may cause the participants to have an influenza vaccine response similar to those that are healthy. Literature examining this possibility is limited, as most studies consider the incidence of secondary acute respiratory illness, peripheral blood mononuclear cell counts, influenza related hospitalization, viral pneumonia, and mortality [13, 19, 27-30]. this is one of the first studies that look at how medications, especially ACEi and ARB, can impact HAIs to influenza vaccination.

Vardeny *et al*. [31] observed that participants with heart failure had statistically significant decreases in HAI titers following influenza vaccination against the H3N2 component when compared to their healthy control participants. There was a non-significant decrease in HAI titers against the H1N1 and B/Victoria components in the vaccine. In a separate study by Vardeny et al, In addition, participants with heart failure had no statistical difference in seroprotection or seroconversion [32]. However, there was a downward trend in low seroconversion among the participants with heart failure when compared to healthy controls (64% vs 82%) [31], (53% vs 88%) [32] across the two studies. Participants with heart failure had no difference in percentage of seroconversion (73% vs 73%), measured at 2-4 weeks post-vaccination [33], but 11-12 month post-vaccination, these same patients had significantly lower (*p* = 0.04) HAI titers compared to the healthy controls (30 vs 60 titers) [33].

Assessing the effect of ACEi and ARBs was not the goal in previous studies, however, most of the participants with heart failure in these trials were taking either an ACEi and ARB [31-33] and it is not clear if the reported results were primarily due to the heart failure condition or due to the administration of these medications. Furthermore, Vardeny *et al*. [31] did not control for the age (57.8 ± 12.9 vs 47 ± 10, *p* = 0.04) of the participants, since older people have lower HAI responses following vaccination [34]. In this study, participants selected for analyses were controlled for age, sex, bmi allowed for a more complete analysis with a lower number of confounders than previous studies [31-33].

This is the first study to analyze the effects of ACEi and ARB medications on HAI responses following influenza vaccination. However, there were three main limitations in this study. First, the presence of other immuno-modulating comorbidities, such as hyperlipidemia and arthritis may have contributed to the inflammatory state of the participants. Second, some of the participants also were taking statin medications, which can have negative impacts on the HAI GMTs [35]. Finally, all participants in this study were older than 65 years of age, which may have played a negative role in seroconversion and HAI responses following influenza vaccination.

Overall, people on these HTN medications had no difference in HAI titers following vaccination compared to healthy control participants with lower seroconversion rates compared to the non-healthy control participants. Further studies are needed to compare the inflammatory markers in people taking ACEi/ARB to healthy people not taking these medications. Furthermore, research on the cell-mediated T- and B-cell responses following influenza vaccination of these participants may add to an improved understanding of the mechanism(s) associated with these medications on HAI seroconversion following influenza vaccinations.

## Data Availability

All data produced in the present work are contained in the manuscript

