## Supplemental Figure 1 for "Influence of ACEi and ARB use on HAI Response to Seasonal Influenza Vaccination": Bunks and Ross Supp 1.pdf

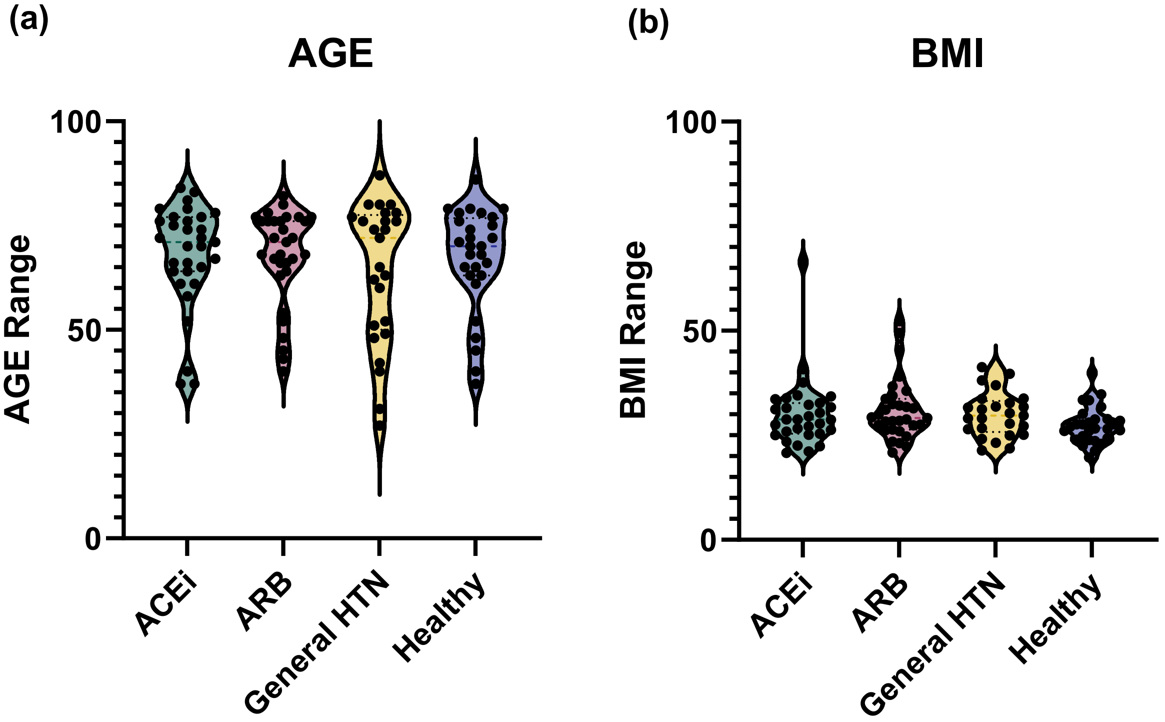


**Supplemental Figure 1**: Violin plot for (a) AGE and (b) BMI across all of the cohorts. Shows that the cohorts were controlled for factors that are established in impacting HAI response.
